# Sample size in social contact surveys for epidemic modelling

**DOI:** 10.64898/2026.03.30.26349407

**Authors:** Ellen Brooks-Pollock, Leon Danon

**Affiliations:** Population Health Sciences, Bristol Medical School, University of Bristol, Bristol BS8 2BN, UK; School of Engineering Mathematics, Jean Golding Institute, University of Bristol, Bristol BS8 1TW, UK

**Keywords:** social contact survey, epidemic model, reproduction number, sample size, infectious disease, contact matrix, POLYMOD, survey methodology

## Abstract

**Background:** Social contact surveys, which measure who-contacts-whom, are widely used to inform infectious disease transmission models and estimate the reproduction number (R), a key metric for assessing epidemic risk. Despite their widespread use, sample size calculations are not routinely performed.

**Aims:** To assess the impact of sample size on estimates of R and determine a practical target sample size for social contact surveys used in epidemic modelling.

**Methods:** We conducted a review of social contact surveys (2008-2025) to characterise current practice. We characterised the impact of survey size on epidemic metrics using two social contact surveys, the UK Social Contact Survey and POLYMOD (Europe) and two methods. For each dataset and approach, we generated repeated subsamples and calculated the resulting reproduction numbers, characterised their distributions and measured uncertainty.

**Results:** We identified 107 unique social contact surveys from 57 studies. Sample sizes ranged from 30 to more than 10,000 participants, with a median of 1,438. One quarter of surveys contained fewer than 1,000 participants. From our simulations, we find that sample sizes below 200 individuals can result in highly variability reproduction numbers. Increasing sample size increases precision, and the most meaningful gains are up to 1,300 individuals. Increasing sample sizes over 3,000 individuals leads to smaller gains.

**Conclusions:** A minimum sample size of approximately 1,200–1,300 participants appears sufficient for general-purpose use. These findings support the inclusion of sample size considerations in the design, reporting and interpretation of social contact surveys used for epidemic intelligence and public health decision-making.

## INTRODUCTION

Social contact surveys have become a crucial data source for estimating real-time epidemic risk in a population and informing structured infectious disease models[1,2]. Prior to the POLYMOD study of age-specific social contact patterns across eight European countries[4], it was often assumed that obtaining accurate empirical data on who-mixes-with-whom at scale would be prohibitively difficult, given ambiguities in defining what constitutes a “contact” and the burden of detailed recording[6]. POLYMOD demonstrated both the feasibility of collecting diary based contact data and the presence of robust, highly structured age-specific mixing patterns that were remarkably consistent across countries, validating the value of such data for transmission modelling and vaccination policy design. Following this, social contact surveys have been implemented in multiple settings and time periods, including during the 2009 influenza and COVID-19 pandemics[7,8], where they were used to measure behavioural change and the impact of non-pharmaceutical interventions on transmission.

Social contact data can be obtained using questionnaire based surveys (paper, online or interviewer administered) or inferred indirectly from demographic, census or household composition data, with each approach involving trade-offs between logistical feasibility, recall bias and the level of detail on contact type and setting[1,9,10]. The most common format for questionnaire based social contact surveys is to ask participants to list their social contacts on the previous day, usually distinguishing between physical and conversational contacts and collecting information on age, setting and duration. Survey instruments have been refined over time to reduce respondent burden and improve data quality, for example by allowing participants to report groups of similar contacts (such as an entire school class) rather than enumerating each individual separately[11,12]. Despite these improvements, completing a social contact diary can remain time-consuming, particularly for individuals with many daily contacts, contributing to non-response, partial completion and dropout in longitudinal designs. Qualitative work has highlighted the importance of participants’ understanding of the survey’s purpose for eliciting accurate responses and sustaining engagement over time[13–15].

Traditional sample size calculations in epidemiological studies typically rely on specifying an expected effect size, the variability of the outcome and the desired power and significance level[16]. However, these approaches are not immediately applicable to social contact surveys, which are often observational, do not test a single, pre-specified hypothesis, and aim instead to characterise complex contact patterns that feed into model-based estimates such as the reproduction number. In addition, social contact surveys frequently face practical constraints, including limited participation rates, survey fatigue and potential bias towards responding on days with fewer contacts[17], all of which complicate the relationship between sample size and the precision of derived epidemiological quantities. As a result, sample sizes in published social contact surveys have typically been determined by pragmatic considerations such as budget, recruitment channels and logistics, rather than by formal power calculations.

Information and data from social contact surveys underpin epidemic analyses, model projections and policy decisions[1,4,18]. Increasing sample size is costly and burdensome for participants, and raises ethical questions if diaries are unnecessarily long or frequent without proper justification. Simple hypothesis-testing approaches to sample size, based on detecting differences in mean contact rates between groups, typically suggest smaller samples and do not reflect how these data are used in transmission models. Here, we seek to determine a practical target sample size for social contact surveys. We first conduct a rapid review of published contact surveys to characterise current practice in participant numbers, then use UK Social Contact Survey [11,12] and POLYMOD to quantify how survey size affects uncertainty in estimates of the reproduction number, and use this to provide a pragmatic estimate.

## AIM

The aims of this study were to review current practice for social contact survey sample size, and to use epidemic metrics to determine a practical target size for social contact surveys used in epidemic modelling.

## METHODS

### Review of current practice

We searched PubMed for all published articles written in English and published between 1 January 2008 and 11 March 2025, using the search string:

((social contact*[Title/Abstract]) AND (survey[Title/Abstract])) AND (infectio*[Title/Abstract]).

Studies were included if they met all of the following criteria: (i) primary reports of social contact data (secondary analyses were excluded); (ii) surveys that included questions specifically addressing the number of social contacts; (iii) studies pertaining to human populations (surveys measuring contact patterns exclusively in animals, or human–animal contact patterns, were excluded); (iv) infectious disease focus and (v) surveys encompassing any age group or demographic.

The first author screened titles and abstracts against the inclusion criteria and reviewed full texts where necessary to determine eligibility. Reference lists of included full texts were searched for additional studies missed by the initial search strategy; these were included if they met the inclusion criteria.

From each included study, we extracted the number of participants and the survey setting or country. Descriptive analyses were conducted on the extracted sample sizes.

### Sensitivity of epidemic measures to social contact sample size

To explore the impact of sample size on epidemic measures, we used data from two commonly-used social contact surveys, and two methods for estimating the reproduction number from social contact data.

#### 1. POLYMOD

POLYMOD was an eight-country European social contact survey conducted in 2005[4]. The surveyed 7,290 individuals on their social contacts for the preceding day. Demographic characteristics of the contacts included age, gender and context. POLYMOD is the most widely used social contact surveys for epidemic modelling[1]. Data are typically incorporated into models by reducing the data to an age-specific contact matrix which is used within an age-structured compartmental model. For the purposes of this study, we used data from all eight countries to create an age-specific mixing matrix in 10-year age bands. We calculated the population reproduction number as proportional to the dominant eigenvalue of the age-specific contact matrix[5,19].

#### 2. UK Social Contact Survey (UKSCS)

UK Social Contact Survey (UKSCS) was conducted in 2010 and surveyed 5,861 individuals using a paper and online survey[11,12]. The UKSCS also asked individuals to record their social contacts on the preceding day, but with additional option to report groups of individuals in order to capture individuals with high numbers of contacts[17]. From the UKSCS, we calculated the sum of the squared individual reproduction numbers, as age of contact was not recorded[18]. This approach captures heterogeneity between individuals.

### Simulating social contact survey sample size

We simulated social contact surveys of varying size by drawing repeated subsamples from the full datasets. Sample sizes ranged from 100 to the maximum number of participants; for each, we drew 200 random samples without replacement. For each simulated sample size, we calculated the mean, range and standard deviation in the resulting reproduction numbers, and the Kolmogorov-Smirnov (KS) statistic as the maximum difference between the cumulative distribution of reproduction numbers compared to a sample of 5,000 individuals.

### Reproducibility and ethics statement

Code was written using R Studio, R version 4.5.3 (2026-03-11). All data used in this article are anonymous and in the public domain, therefore no ethical approval was required to conduct this study. POLYMOD data are available from https://socialcontactdata.org/ and the UKSCS data from http://wrap.warwick.ac.uk/54273/. Processed data and code are available at github.com/bristol-vaccine-centre/contact-sample-size.

## RESULTS

The initial search for social contact surveys related to infectious disease transmission returned 145 records. Following title and abstract screening, 93 studies were taken forward for full-text review. The most common reasons for exclusion were not a contact survey (n=28), not related to infectious disease transmission (n=23) and secondary analyses (32). A further two studies were identified through reference list searching, giving 95 studies assessed for review. After full-text review, 57 studies were included in the final analysis. Because several papers reported more than one survey or setting, these 57 studies corresponded to 107 unique social contact surveys.

There was a wide range in reported sample sizes across the 107 surveys (Figure 1). The smallest was a pilot study of 30 participants, while the largest exceeded 10,000 — five surveys of this scale had been embedded within larger national studies, such as a census. Smaller surveys tended to focus on specific subgroups, such as university students. One quarter of all surveys contained fewer than 1,000 participants, and three-quarters contained fewer than 2,500 participants. The median sample size was 1,438 participants.

**Figure 1.**
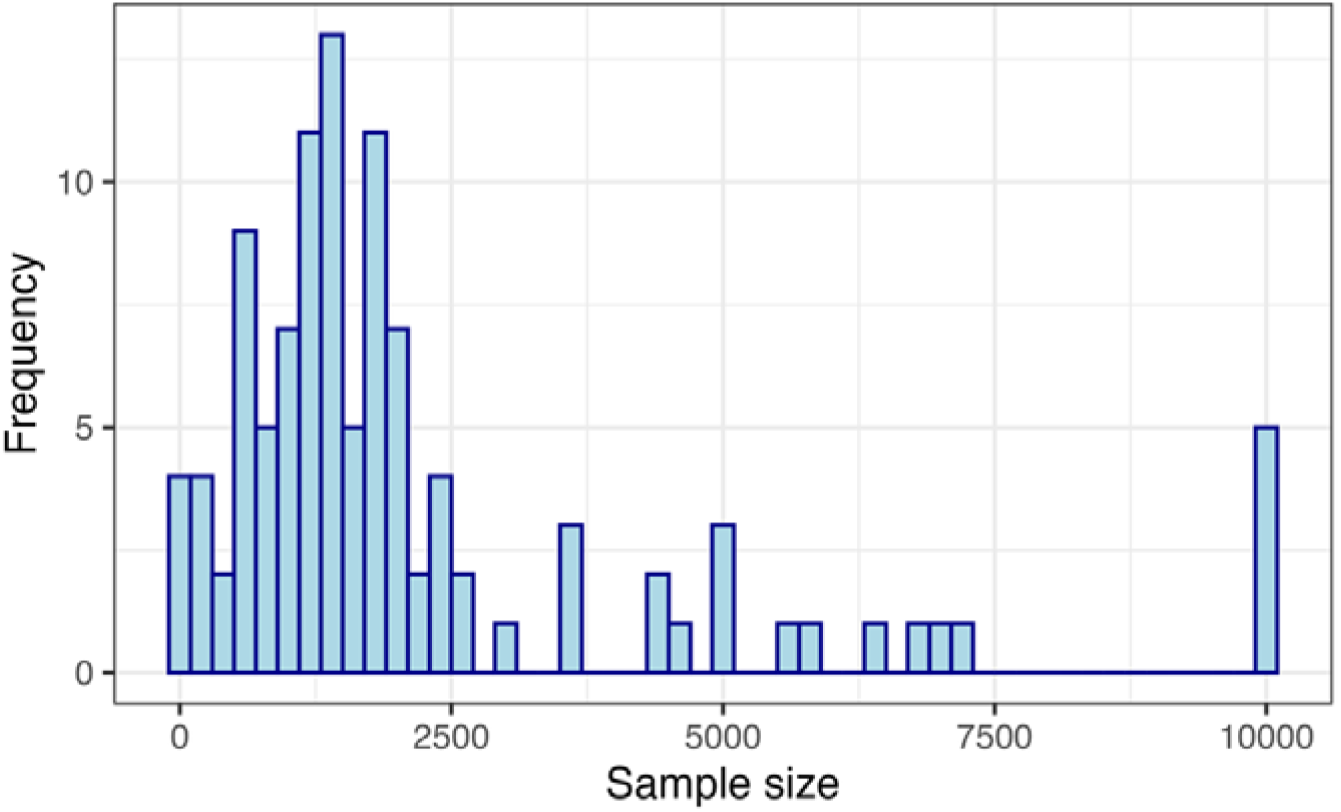
Distribution of social contact survey sizes from a rapid review of 107 surveys.

### Impact of sample size on estimates of the reproduction number

To assess how social contact survey sample size influences epidemic metrics, we analysed two widely used datasets and applied two methods to estimate the population-level reproduction number. Across both datasets and methods, precision in the reproduction number increased with sample size (Fig. 2; Table 1). Estimates derived using the dominant eigenvalue approach with POLYMOD data showed consistently lower uncertainty than those obtained using the sum of squared individual reproduction numbers (R^2^) with UKSCS data.

**Table 1:**
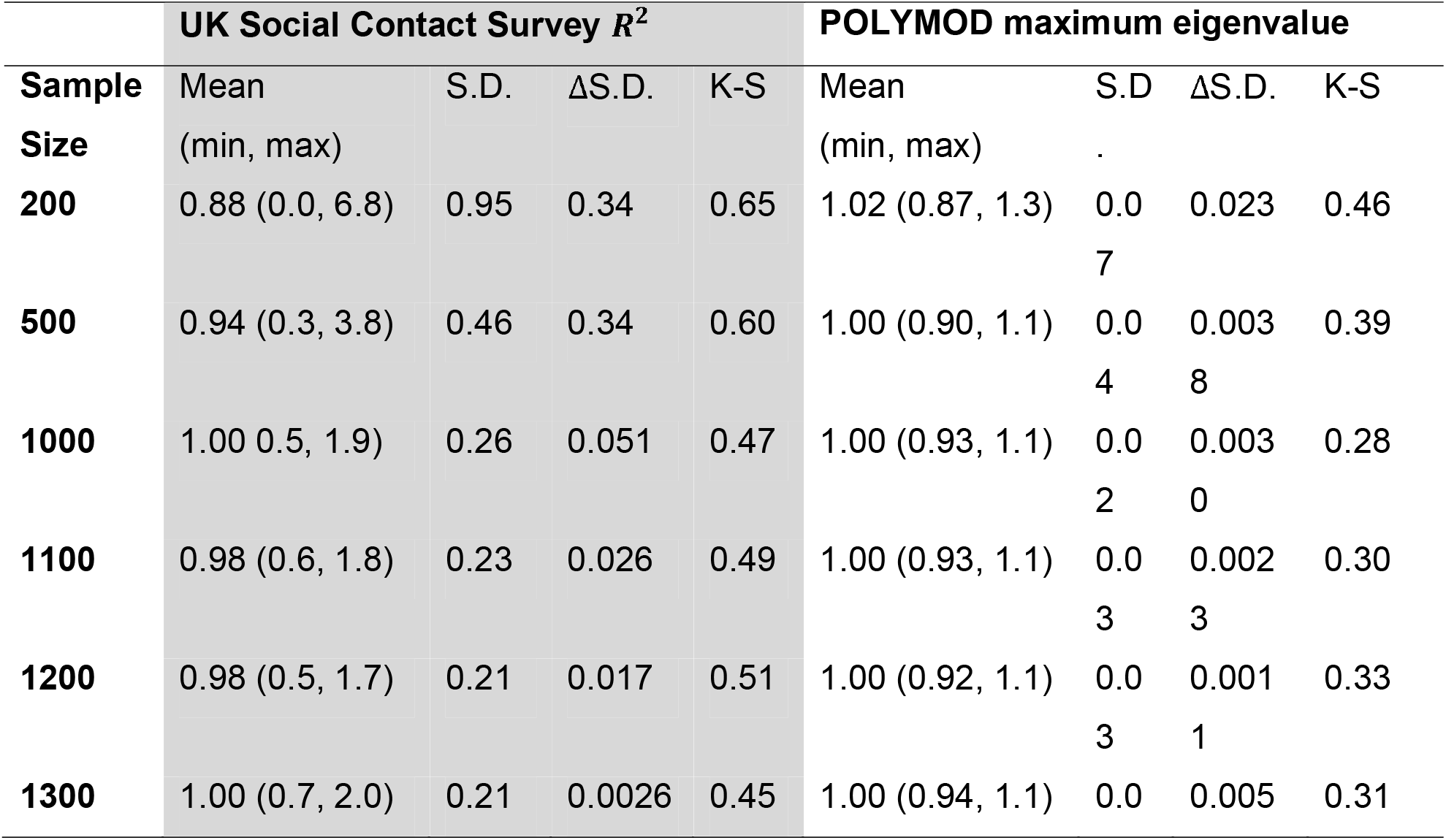
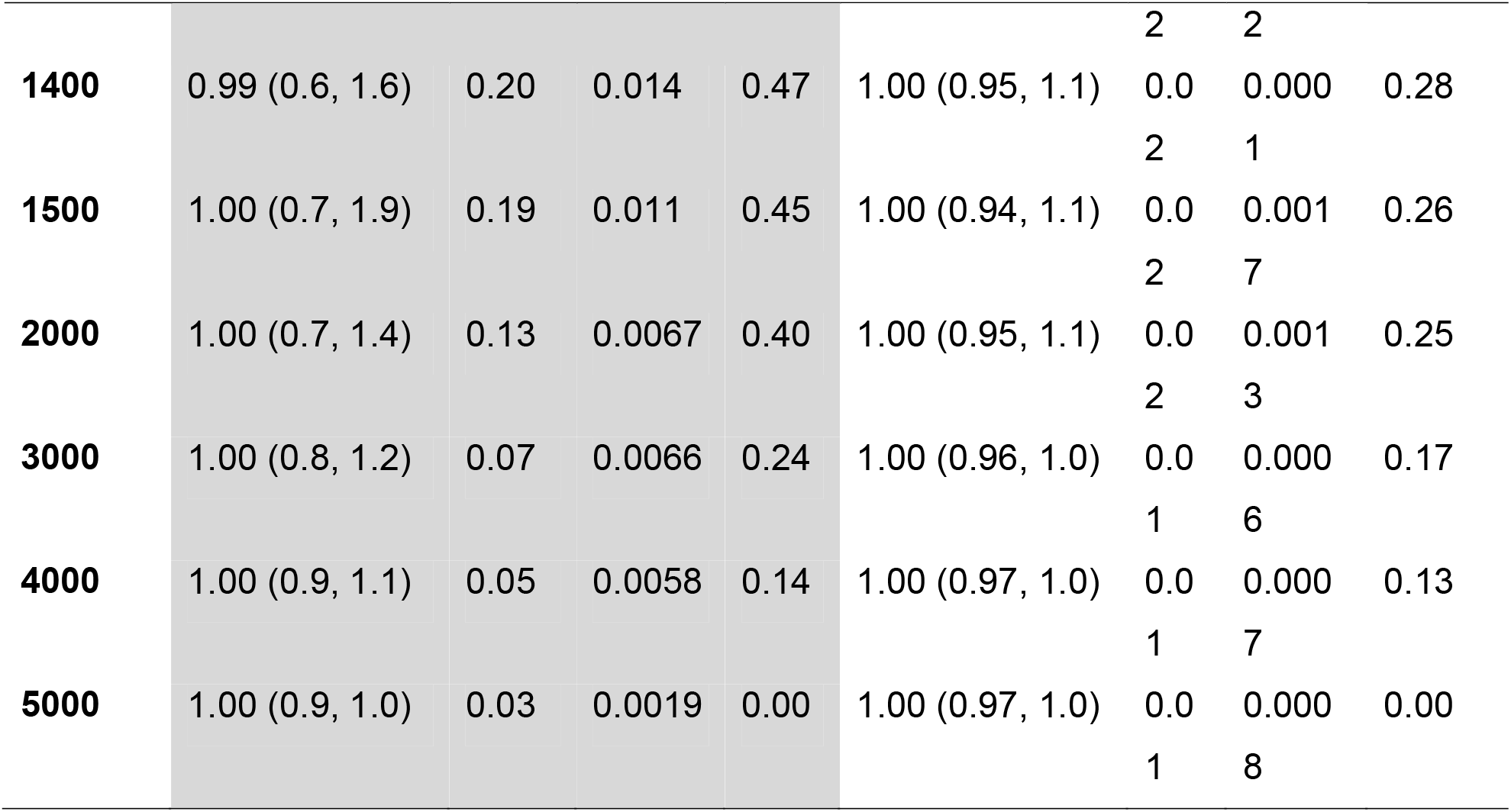
Statistics of the estimated reproduction number (normalised to 1) as a function of social contact survey sample size for two social contact surveys and two methods for calculating the reproduction number. Values are reported to 2 significant figures where appropriate.

**Figure 2.**
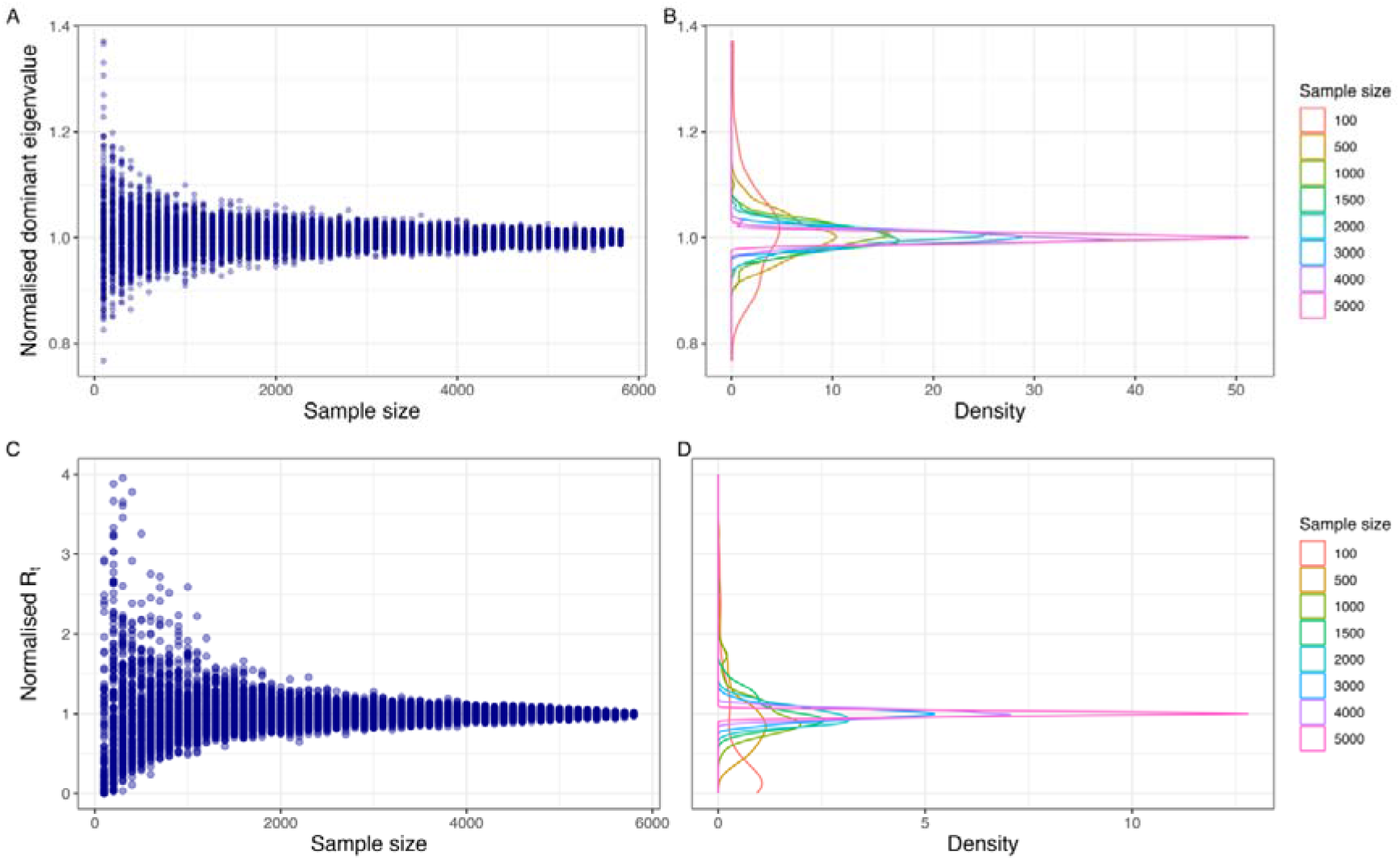
Relationship between number of participants contributing social contact data (sample size) and estimated reproduction number. Panels A and B show the normalised dominant eigenvalue of the next generation matrix derived from the POLYMOD study; panels C and D represent the normalised reproduction number derived from the UKSCS. The points in Panels A and C show individual values derived from a single sample and panels B and D summarise the distributions at specific values of the sample size shown as different colours.

At sample sizes below 200 individuals, the dominant eigenvalue method produced R estimates ranging from 0.87 to 1.3 (2 s.f.), whereas the R^2^ approach led to a wider range (0 to 6.8; 2 s.f.). The Kolmogorov–Smirnov (KS) statistic—representing the maximum distance between cumulative distributions—exceeded 0.5 for sample sizes below 1,200 under the R^2^ approach, and 0.3 for sample sizes below 1,100 under the dominant eigenvalue approach, suggesting larger differences between resulting R estimates.

For both methods, the relative reduction in standard deviation decreased with increasing sample size up to approximately 1,300 individuals, after which gains in precision were smaller. Above 3,000 individuals there were small differences in resulting estimates.

## DISCUSSION

We found considerable variability in the number of participants included in published social contact surveys, ranging from 30 to over 10,000 individuals, with a median sample size of 1,438. A quarter of surveys containing fewer than 1,000 participants. We show that small social contact surveys risk resulting in inaccurate estimates of epidemic risk assessment, and that very large surveys might be sampling unnecessarily. As a guide, sampling a minimum of 1,300 individuals increases confidence in the derivative results.

Sample size estimates have been largely absent from the design of social contact surveys, despite their direct consequences for the reliability of transmission models. Existing systematic reviews have catalogued the diversity of survey methodologies and settings[1,2], but none have addressed the statistical adequacy of the samples collected. Our findings provide a quantitative basis for evaluating published surveys and for measuring the uncertainty in epidemiological estimates. In particular, surveys containing fewer than 1,000 participants should be used with caution to parameterise transmission models. Epidemic forecasts derived from contact survey data should be interpreted in the context of the sample size of the underlying survey, and we recommend that sample size be reported as a standard element of contact survey publications.

The rapid review of social contact surveys provides a snapshot of current practice; however, it was restricted to English-language publications indexed in PubMed, and surveys reported in other languages or in grey literature were not captured. We used the population-level mean reproduction number as our primary outcome metric; other quantities derived from contact data such as the critical vaccination threshold or age-specific incidence rates may show different sensitivity to sample size. Data quality represents a further constraint that is independent of sample size: survey fatigue, underreporting of high-contact days, and the systematic underrepresentation of certain demographic groups are well-documented in contact surveys[13,15,17,20], and affect the validity of derived estimates in ways that cannot be remedied by increasing recruitment alone.

Our analysis was conducted using two social contact surveys from the UK and Europe. While the two surveys yielded broadly similar results, the differences illustrate the impact of survey design and methodology. Social contact degree distributions are often heavy-tailed, i.e. with highly connected individuals occurring more often than might be expected. Sampling heavy-tailed networks can lead to underestimating the variance; a study of heavy-tailed sexual networks suggested that sampling should be designed to capture this variability[21]. We observed more variability in the UKSCS than POLYMOD, due to the facility to record groups in the UKSCS, and the mixing matrix approach, mostly used with POLYMOD-style data, which averages over age bands. We found that for 10-year age bands, a sample size of 1,100 individuals was sufficient; for finer grained models larger sample sizes are required.

Our findings have implications for pandemic preparedness and response across Europe. Social contact data provide a leading indicator of the reproduction number, as they are more closely linked to transmission events than lagged indicators such as diagnoses, hospital admissions or deaths. During the COVID-19 pandemic, social contact data were used to estimate the impact of physical distancing restrictions on transmission[8]. Our results highlight the vulnerability of such early estimates to study design and sample size and suggest that further work is needed to tailor sampling schemes to capture highly connected individuals. Establishing a minimum sample size for social contact surveys will improve comparability between settings, supporting more consistent epidemic intelligence. This study provides the first exploration of sample size and R uncertainty, enabling more reliable estimation of R and strengthening the evidence base for coordinated public health decision-making.

## Supporting information

PRISMA flowchart

## Data Availability

All data produced are available online at https://github.com/bristol-vaccine-centre/contact-sample-size

https://socialcontactdata.org/

http://wrap.warwick.ac.uk/54273/

