## Supplementary material for "Sample size in social contact surveys for epidemic modelling": PRISMA flowchart

**Identification of studies via other methods**

**Identification of studies via databases**

Records identified from:

Citation searching (n = 2)

Records removed *before screening* (n=0)

Records identified from PubMed (n=145)

**Identification**

Records excluded (n = 52)

Not a contact survey (n=17)

Not human (n=3)

Not infectious disease (n=18)

Secondary analysis (n=13)

Duplicate (n=1)

Title and abstract screening

(n = 145)

Reports not retrieved

(n = 0)

Title and abstracts screened

(n = 2)

**Screening**

Reports excluded (n=38)

Duplicate (n=1)

Not human (n=1)

Protocol development (n=1)

Not infectious disease (n=5)

Not a contact survey (n=11)

Secondary analysis (n=19)

Text screening

(n = 93)

Reports/manuscripts included in review

(n = 57)

Surveys included

(n = 107)

**Included**

*Adapted from:*  Page MJ, McKenzie JE, Bossuyt PM, Boutron I, Hoffmann TC, Mulrow CD, et al. The PRISMA 2020 statement: an updated guideline for reporting systematic reviews. BMJ 2021;372:n71. doi: 10.1136/bmj.n71. For more information, visit: <http://www.prisma-statement.org/>
